# Integration of Kalman filter in the epidemiological model: a robust approach to predict COVID-19 outbreak in Bangladesh

**DOI:** 10.1101/2020.10.14.20212878

**Authors:** Md. Shariful Islam, Md. Enamul Hoque, Mohammad Ruhul Amin

**Affiliations:** Department of Mathematics and Physics, North South University, Dhaka, Bangladesh; Computational Physics Group, Department of Physics, Shahjalal University of Science and Technology, Sylhet - 3114, Bangladesh; Computer and Information Science, Fordham University, New York, USA

## Abstract

As one of the most densely populated countries in the world, Bangladesh have been trying to contain the impact of a pandemic like COVID-19 since March, 2020. Although government announced an array of restricted measures to slow down the diffusion in the beginning of the pandemic, the lockdown has been lifted gradually by reopening all the industries, markets and offices with a notable exception of educational institutes. As the physical geography of Bangladesh is highly variable across the largest delta, the population of different regions and their lifestyle also differ in the country. Thus, to get the real scenario of the current pandemic across Bangladesh, it is essential to analyze the transmission dynamics over the individual districts. In this article, we propose to integrate the Unscented Kalman Filter (UKF) with classic SIRD model to explain the epidemic evolution of individual districts in the country. We show that UKF-SIRD model results in a robust prediction of the transmission dynamics for 1-4 months. Then we apply the robust UKF-SIRD model over different regions in Bangladesh to estimates the course of the epidemic. Our analysis demonstrate that in addition to the densely populated areas, industrial areas and popular tourist spots are in the risk of higher COVID-19 transmission. In the light of these outcomes, we provide a set of suggestions to contain the pandemic in Bangladesh. All the data and relevant codebase is available at https://mjonyh.github.io.

**Highlights:**

- We integrate the UKF with classic SIRD model for the better estimation of the COVID-19 diffusion of 64 districts in Bangladesh.
- Nationwide analysis show the strong correlation between population density and the number of COVID-19 positive cases in the country.
- Industrial zones and popular tourists spots are at greater risk of spreading the Coronavirus.
- With the better assessment of the COVID-19 cases dynamics, the Government will find effective policies to contain the current pandemic.

## 1 Introduction

The world has been braving the COVID-19 crisis by which more than 52 million people across 113 countries over the globe got infected as of *November* 11, 2020. The Corona virus disease was first identified in Wuhan, the capital of Central China’s Hubei province, on December 2019. The World Health Organization (WHO) declared it as the public health emergency on January 20, 2020 and later as Pandemic on March 11, 2020 [1, 2]. Although China, Italy, Spain, USA and UK were severely affected by COVID-19 at first, beginning from April 2020 onward, countries from Global South are also impacted by this devastating Corona virus. As of *August* 30, 2020, the lower middle income country Bangladesh has the second lowest number of tests conducted per confirmed cased of COVID-19.

The novel Corona virus can be transmitted via three commonly accepted modes, namely direct contact with an infected person, contaminated objects, and larger respiratory droplets or small airborne droplets [3]. Usually the virus infected person shows symptoms like fever, shortness in breath, cough, diarrhea, pneumonia and loss of smell, whereas the corona positive patient with premedical conditions such as diabetics, pneumonia and high blood pressure might experience the fatal death if proper medical treatment cannot be sought for in time [4, 5]. Since there are no recommended medicines or remedies available for treating COVID-19 disease as of now, many countries are setting up an array of measures including wearing face mask, early detection of infected people, contact tracing, isolation of infected people, increasing the number of testing per day, and imposing lockdown to curb the COVID-19 outbreak [6, 7]. Recent studies also support the effectiveness of wearing face mask, social distancing, and imposing lockdown on the declining of both infection and death rate [8].

Bangladesh is one of the most densely populated (ranked 8^*th*^ worldwide) and under developing countries in the world. The recent economic development of this country makes it very competent in the South Asian region, especially in terms of foreign exchange reserves, reaching as high as second position in South Asia [9]. The first COVID-19 positive case was identified in Bangladesh on March 8, 2020. The Government of Bangladesh imposed a set of strict measures and closed all educational institutions, government, non-government offices including garments and factories [10] on March 26th, 2020. As a result, the number of infection and fatality rates are less compared to India and Pakistan. In Bangladesh, the contamination numbers spilled over 425,353 confirmed cases with death toll of more than 6,127 and it is still growing each day [11].

To understand the dynamics of a pandemic, mathematical model can play a major role. The classic mean-field Susceptible-Infected-Recovery-Death (SIRD) model by Kermack and McKendrick [10, 12], is frequently used to illustrate a quantitative picture of COVID-19 outbreak for many countries [6]. However, spread of an epidemic is a sophisticated process that depends largely on the mutated strains of the virus and its principle vectors [13, 14]. Moreover, the recorded data of infected, recovered and death cases might miss significant amount of information due to many kinds of biases [15]. For instance, time series of recovered numbers are heavily unreliable as it is solely based on how a country traces its asymptomatic or mildly symptomatic patients. Therefore, usual mean-field, compartment-based models and stochastic spatial epidemic models might estimate the dynamics of an epidemic with higher error margin [16–19].

The SIRD model is associated with a set of differential equations which compute values for an instantaneous event, whereas the time series of COVID-19 cases that we observe everyday is a discrete event. Therefore, all the daily events of COVID-19 cases lack the instantaneous effect in the differential equation based SIRD model. Here, we use the prediction based Unscented Kalman Filter (UKF) model to derive the dynamics. UKF is a classic non-linear estimation algorithm that accurately and timely forecasts the dynamic state in a nonlinear system. UKF has been used in various areas such as, navigation, target tracking, structural dynamics and vehicle positioning due to its high accuracy and rapid convergence merits [20, 21].

In this study, we integrate the UKF with classic SIRD model to examine the underlying process of COVID-19 transmission in a more precise way. As Bangladesh has 64 districts which are uneven in terms of population density, economical significance and geographical locations, we estimate the transmission trend of 64 districts to evaluate the complete picture of the outbreak in Bangladesh.

## 2 Method

We used the standard SIRD model to estimate the number of COVID-19 active cases, recovered cases and death cases in Bangladesh. A homogeneous immunity has been considered for the whole country with no transmission from animals and no significant difference between natural death and birth. We segmented the total population size, *N*, into four stages of disease: *S*, susceptible; *I*, infected; *R*, recovered; *D*, death and can be written by *N* = *S* + *I* + *R* + *D* [10, 12, 22, 23]. The interaction between these four stages is illustrated in the Figure 1.

**Fig 1.**
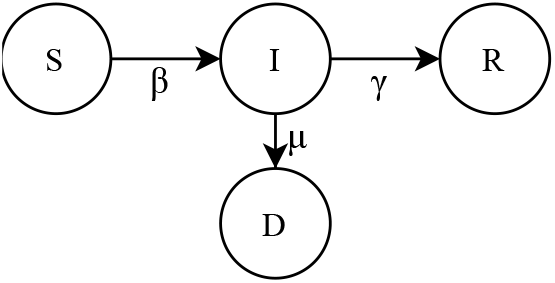
Schematic diagram of the compartmental SIRD model. It represents the total populations (*N*) into four different health status, such as, susceptible (*S*), infected (*I*), recovered (*R*) and death (*D*). The *β, γ* and *µ* denote the rate of infection, rate of recovery and rate of death, respectively.

The differential equations of SIRD model can be written as:

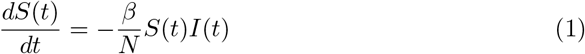

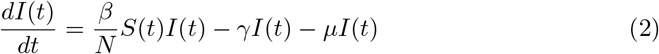

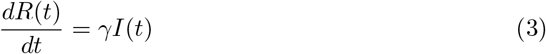

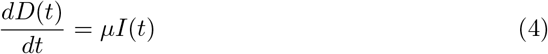

Where *t* denotes the time duration, *S*(*t*) represents the number of susceptible people at time *t, I*(*t*) shows the number of people infected at time *t, R*(*t*) stands for the number of people who have recovered from the infection and *D*(*t*) indicates the number of deaths. The population, *N* = *S*(*t*) + *I*(*t*) + *R*(*t*) + *D*(*t*), is a conserved quantity for every time step [24, 25]. The constant *β, γ* and *µ* illustrate the rate of infection, recovery and deaths, respectively.

The Eq. 1 - 4 have been discretized by forward Euler method as,

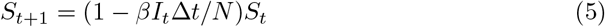

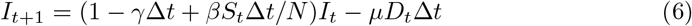

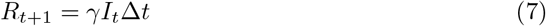

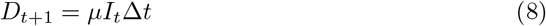

An augmented state vector *X* is presented for simplicity as,

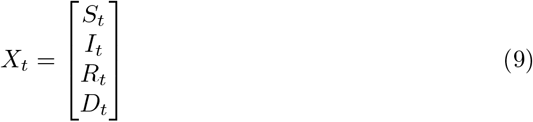

and the discrete-time augmented SIRD model, Eq. 5 - 8, can be written as,

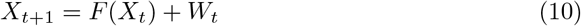

where *F* is the nonlinear term and *W* is the zero mean Gaussian uncertainty with covariance *Q*_*f*_. All types of reported cases, the cumulative confirmed, recovered, deaths and active cases, are incorporated with the model using the vector as,

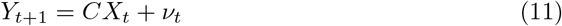

where *ν* is the uncertainties due to the SERS-CoV-2 test results. The considered zero mean Gaussian uncertainty has the known covariance *R*_*f*_.

Let us consider an estimated vector state 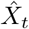 for the Unscented Kalman Filter (UKF), then,

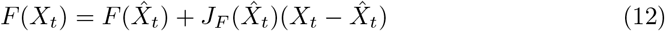

where 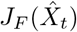 is the Jacobian matrix of *F*(*X*_*t*_), given by,

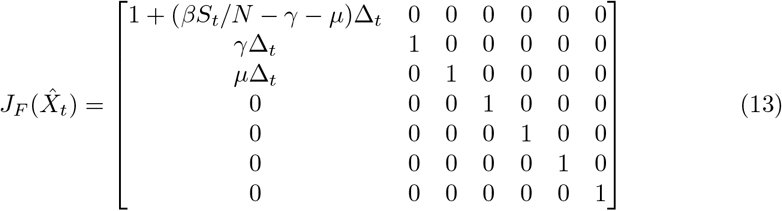

Hence the algorithm of UKF is given by,

### Predict

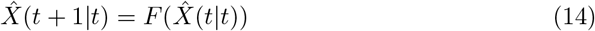

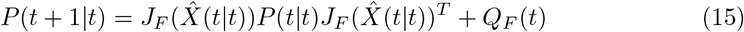

### Update

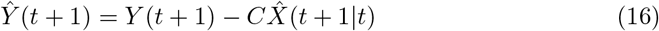

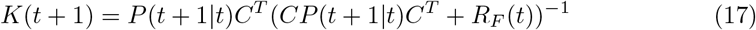

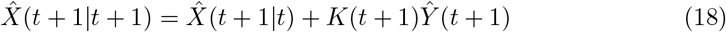

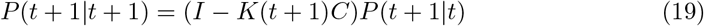

Here, *P* (*t* |*t*) and *C* represent the posterior estimate covariance matrix and the data augmented matrix respectively. We integrated this updated formula of UKF in SIRD model to estimate the dynamics of the SERS-CoV-2 in Bangladesh.

The dynamical behaviour of the class of infected people is described by the basic (*R*_0_), effective (*R*_*e*_) and time-varying (*R*_*t*_) reproduction number. In the case of SIRD model, this are defined as,

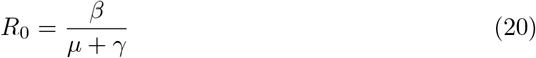

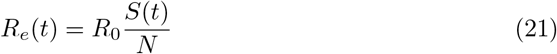

Using Eq. 2, one can find that *S*(*t*) = *N/R*_0_ and *R*_*e*_ = 1, at *I* = *I*_*max*_. Besides, one can estimate the onset confirmed (*C*^*′*^) cases as, 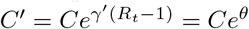, where *θ* = *γ*^*′*^(*R*_*t*_ *−* 1) observes random walk and *γ*^*′*^ vary independently [26]. Thus,

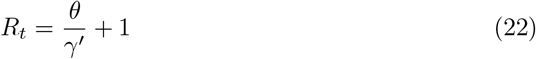

If this reproduction number becomes larger than the ratio between the total population and initial susceptible people, there will be a proper epidemic outbreak [27]. Otherwise, the disease will never lead to a proper outbreak.

## 3 Results

As a state of art, we first evaluate whether UKF-SIRD model provides more robust estimation of COVID-19 active cases in Bangladesh compared to classic SIRD model prediction. To illustrate the active cases of the outbreak, the UKF-SIRD and SIRD model simulations were performed in the GNU octave (Version 5.2.0) and Python programming language environment. It is clearly noticeable in figure 2 that UKF-SIRD model delivers more accurate data estimation compared to normal SIRD model prediction. For instance, the UKF-SIRD model predicts around 85,000 corona active cases at 100 days (third week of June, 2020) relative to 90,000 real cases, whereas SIRD model estimates 120,000 active cases at the same time in Bangladesh. Therefore, these results illustrate the fact that integrated UKF in SIRD model is a robust method which is capable of exclusive prediction of COVID-19 transmission in Bangladesh.

**Fig 2.**
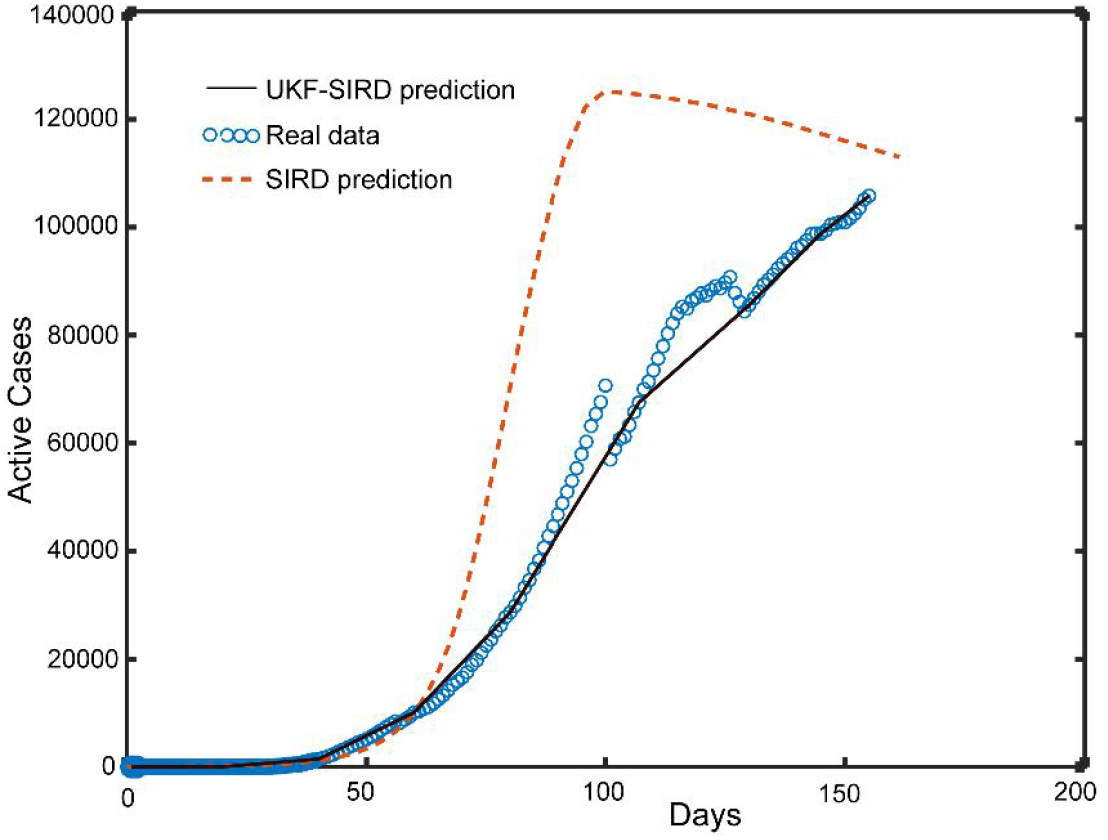
The estimation of corona active cases in Bangladesh by UKF-SIRD and SIRD model. Dynamics of the active cases population has been shown as a function of time. The real data are fitted from March 8 to July 28, 2020.

Bangladesh has 64 districts that are heterogeneous in the context of different population densities, socioeconomic activities and geographical parameters as shown in figure 3. For example, Dhaka, the capital of Bangladesh, has a population density of 23,234 people/sq. km compared to Bandarban (87 people/sq. km) district in Bangladesh. On the other hand, Narayanganj has more industries than Bogra district. Recently, Nadjat and colleagues showed the positive impact of population density on COVID-19 diffusion in Algeria [32]. Thus, considering the heterogeneous density in Bangladesh, general estimation of the COVID-19 dynamics will never match with the real outbreak scenario unless it is done on the district level. To evaluate the local transmission in Bangladesh, next we cluster whole Bangladesh into three groups according to its population density, industrial area and geographical variance.

**Fig 3.**
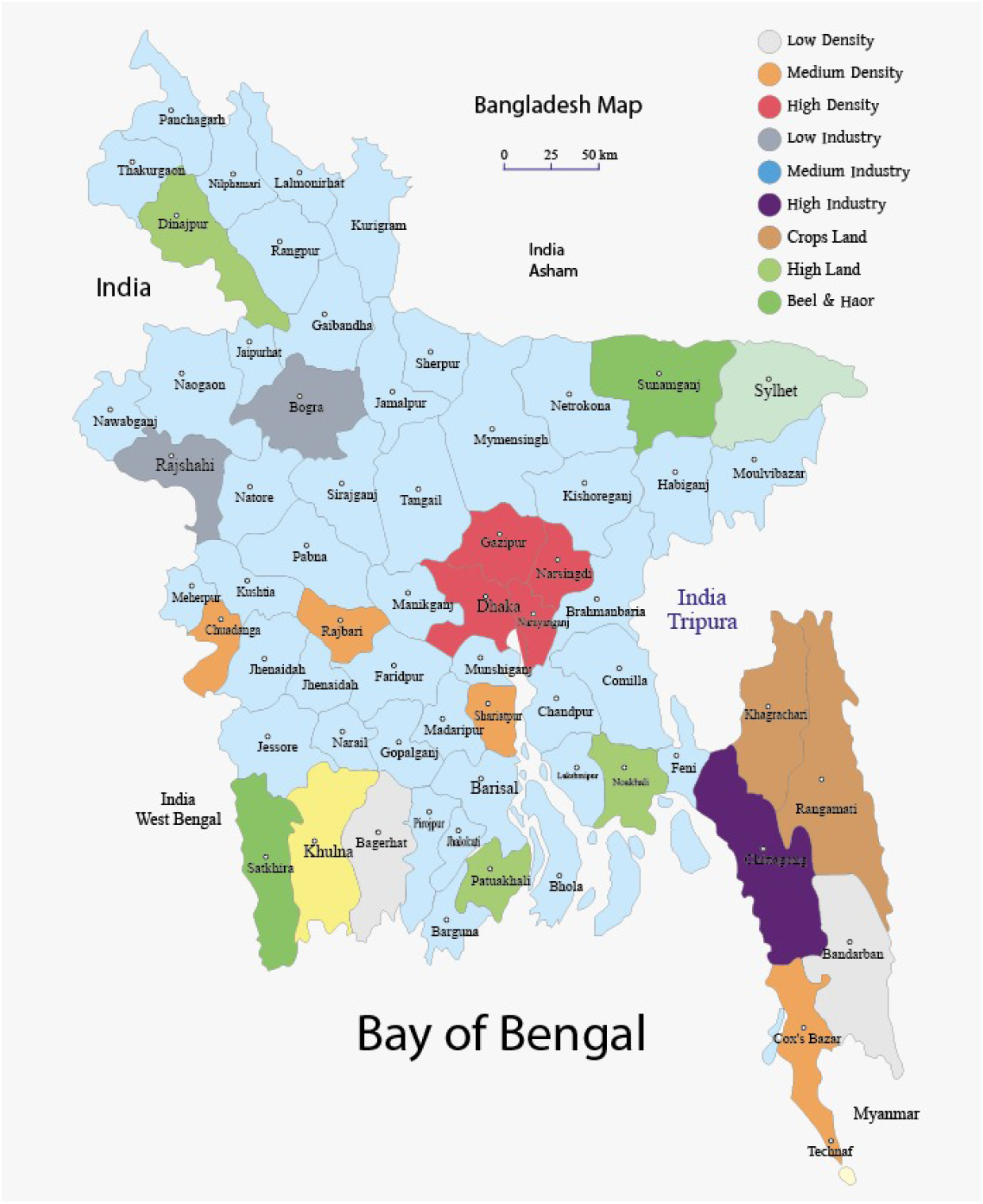
Classification of Bangladesh map based on population density, industrial regions, agricultural land, high land, Beel and Haor

**Fig 4.**
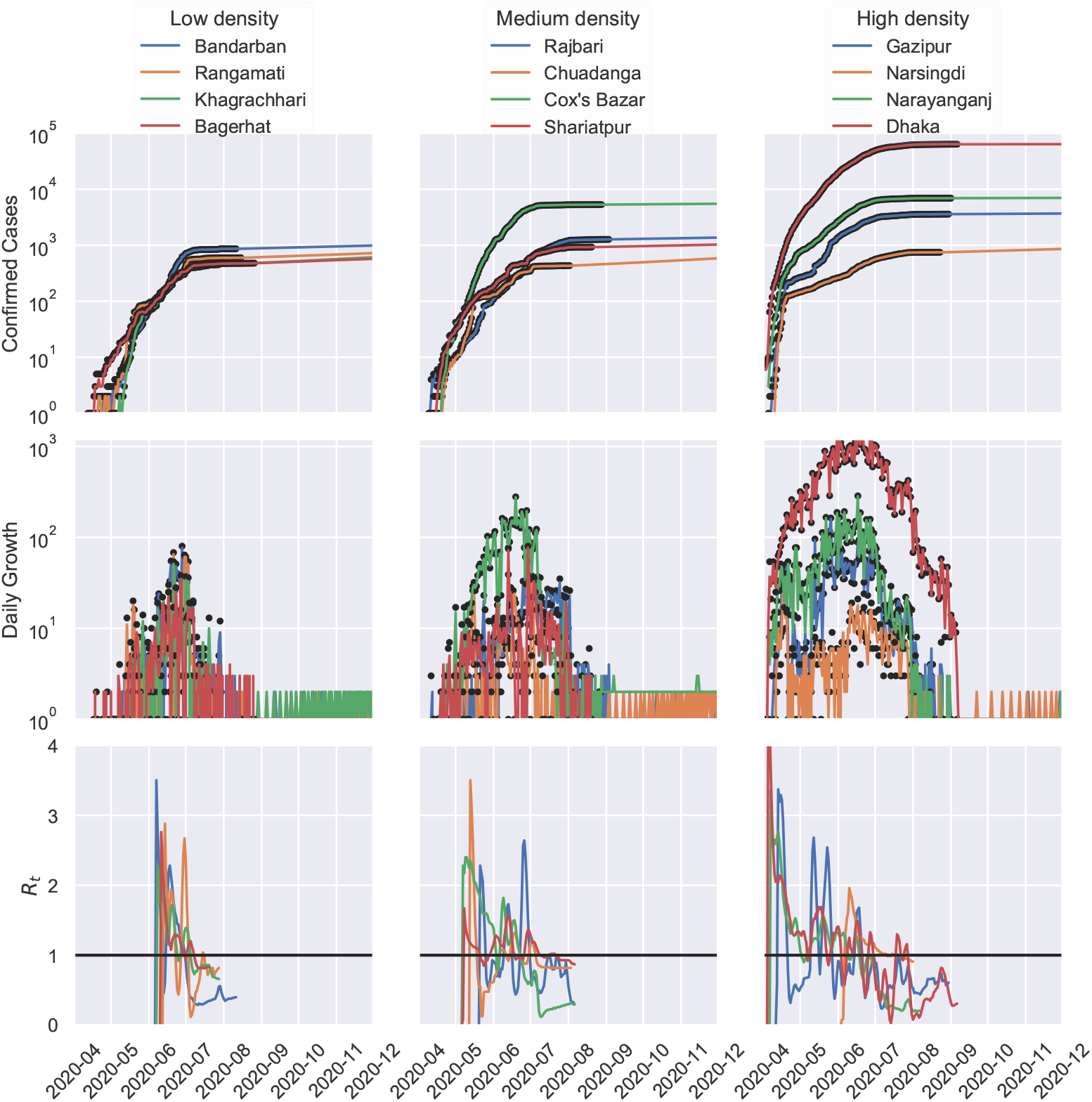
Epidemic evolution of COVID-19 based on confirmed cases, daily growth and *R*_*t*_ values in Bangladesh. The UKF-SIRD model illustrates the positive impact of population densities on COVID-19 confirmed cases and daily growth in major districts such as, Dhaka, Gazipur and Narayanganj.

**Fig 5.**
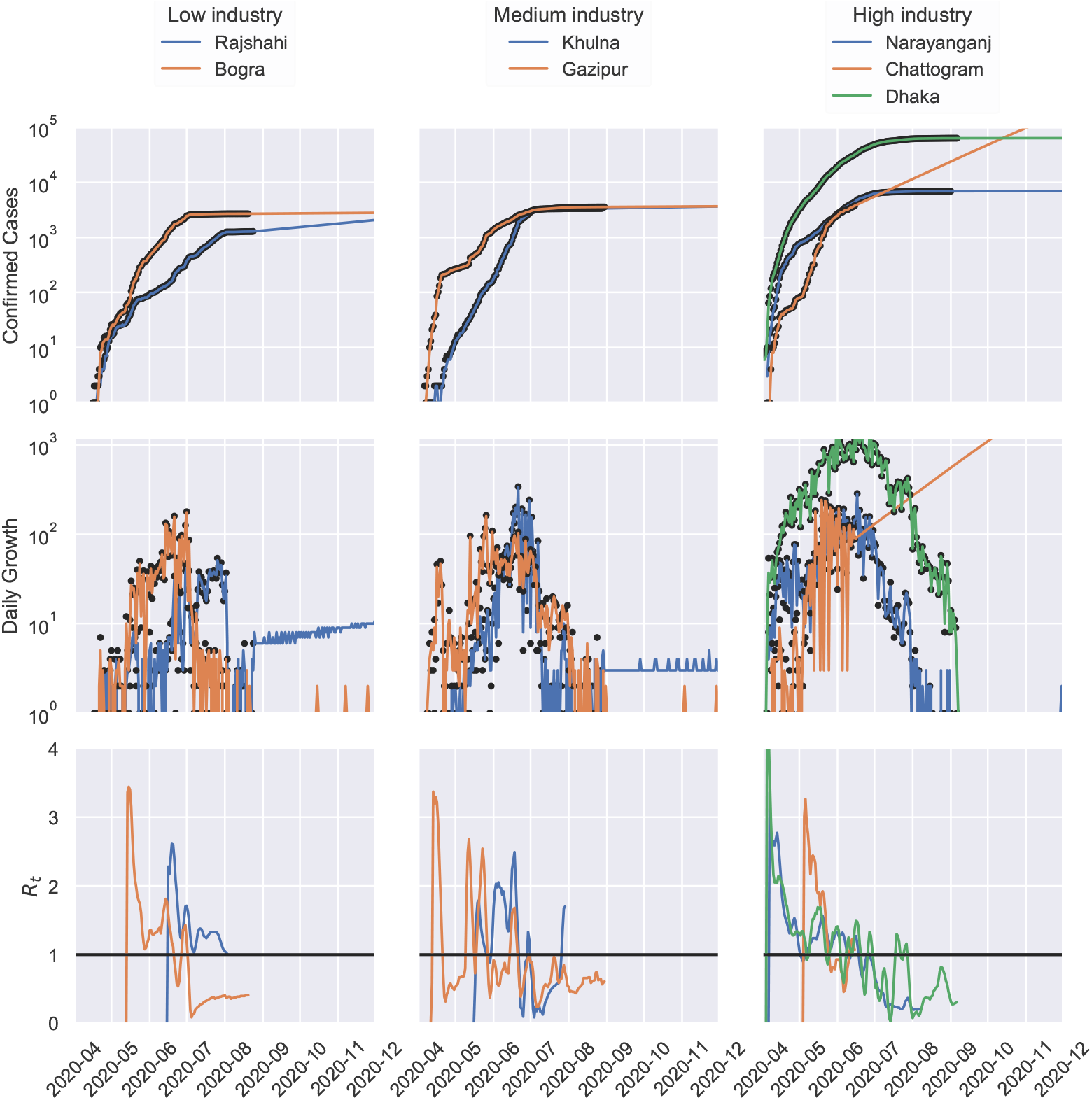
Epidemic evolution of COVID-19 based on confirmed cases, daily growth and *R*_*t*_ values in Bangladesh. The UKF-SIRD model illustrates the positive impact of industrial zones on COVID-19 confirmed cases and daily growth.High Industrial districts such as Dhaka, Chattogram are at high risk for COVID-19 diffusion compared to low industrial area like, Bogra.

**Fig 6.**
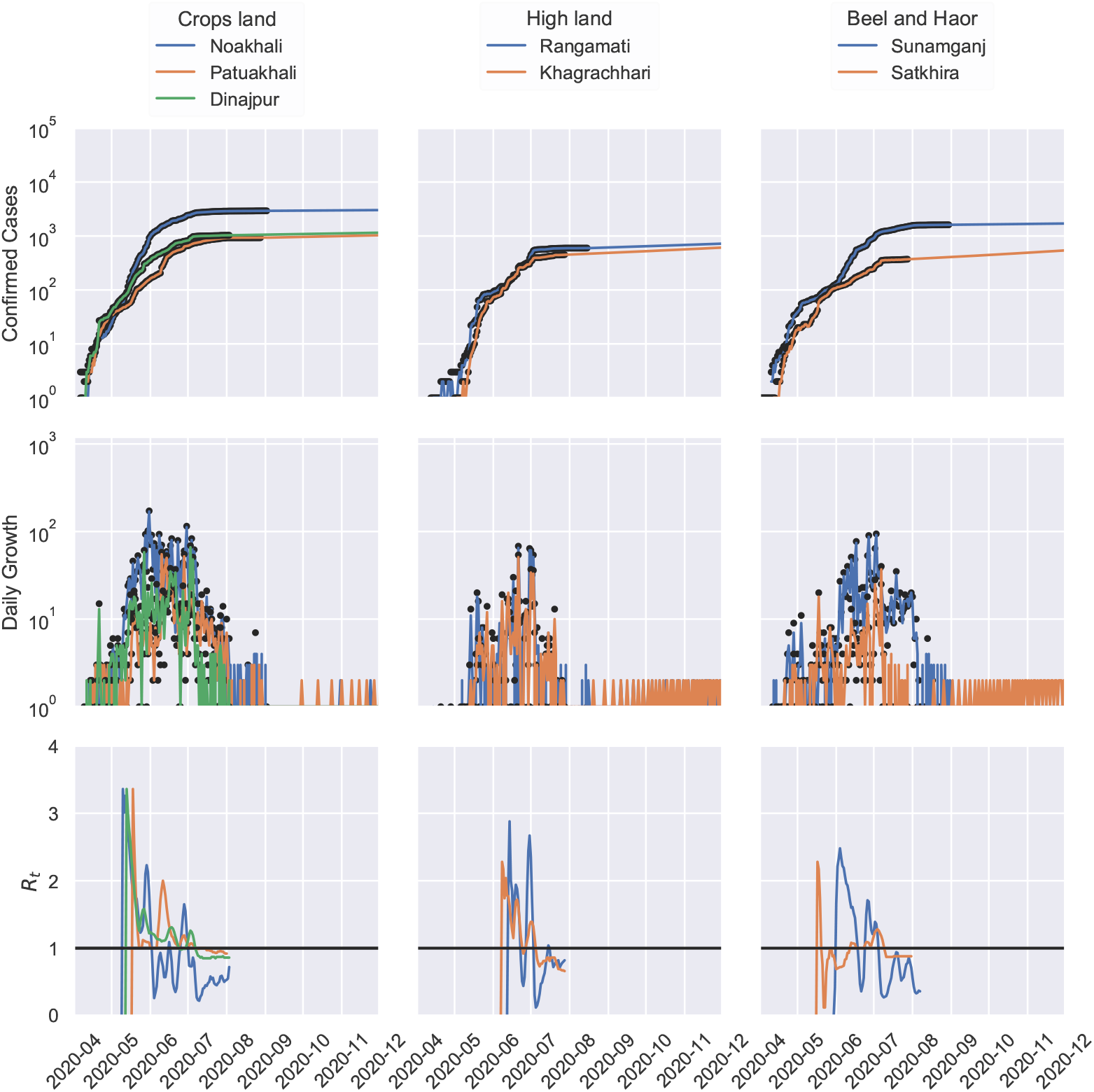
Epidemic evolution of COVID-19 based on confirmed cases, daily growth and *R*_*t*_ values in Bangladesh. The UKF-SIRD model illustrates that Beel, Haor areas (Sunamganj, Satkhira), Highland districts (Rangamati, Khagrachhari) and argicultural zones (Noakhali, Dinajpur) are at low risk for COVID-19 transmission in Bangladesh.

### 3.1 Estimation of COVID-19 diffusion in highly dense cities of Bangladesh

In this article, we consider 12 districts (high to low dense areas) of Bangladesh to examine the spread of the Corona virus infection. Fig. 3.1 clearly shows that densely populated area, Dhaka is severely affected by COVID-19 outbreak as the confirmed cases reach ∼ 100, 000, whereas low dense area, Bandarban has only ∼ 1, 000 positive cases. In addition, the daily growth rate also follows a high trajectory trend for Dhaka compared to Narsingdi, Shariatpur and Bagerhat, as shown in the Fig. 3.1. The *R*_*t*_ values of Dhaka were over 1.0 starting from April, 2020 to end of July,2020, whereas Bagerhat had the *R*_*t*_ values *>* 1.0 for only 30 days (June,2020). Interestingly, we notice that the district of Cox’s Bazar has similar number of corona positive cases ∼ 10, 000 like highly dense district of Gazipur despite the fact that Cox’s Bazar is a moderately dense area in Bangladesh. Since Cox’s Bazar is the most popular tourists spot in Bangladesh, we hypothesize that high mobility in this area might cause the high number of corona positive cases. Strikingly, at the end of July, 2020 *R*_*t*_ value is approaching to *<* 1.0 for all twelve districts in Fig. 3.1, which could lead to the end of this pandemic if the current trend continues for the next 3-4 months.

### 3.2 COVID-19 transmission trend in the industrial areas of Bangladesh

The year 2020 has seen the unprecedented downfall of the economy for both the national and global market, trade, and investment to reduce the infection rate of the novel Corona virus by ceasing the means of transportation and forcing the lockdown measures that shutdown many business organizations. Moreover, many garment factories were temporarily closed to avoid the risk of Corona virus outbreak. Despite the government-announced shutdown to prevent the COVID-19 diffusion in Bangladesh, industrialists reopened the factories for the economic salvation.

However, health experts have shown their grave concerns about the proper guidelines for social distancing inside the local factories, as several ready made garment (RMG) workers have been tested Corona virus positive in the recent days. Therefore, it is important to determine the COVID-19 transmission trend in these industrial districts such as, Narayanganj, Chattogram, Khulna and Rajshahi. Our systematic UKF-SIRD model analysis illustrates a sharp daily growth of corona positive cases for the industrial area, Chattogram, compared to another industrial zone, Narayanganj. Moreover, small industrial region like Rajshahi has less number of cases (*∼* 1, 000) relative to Narayanganj (*∼* 10, 000) and Dhaka (*∼* 100, 000). Furthermore, we observe the uncertain values of *R*_*t*_ for Gazipur district, whereas Khulna show a growing trend of *R*_*t*_ from August, 2020. These analysis clearly forecast the fact that high industrial areas are at greater risk of Corona virus infection in Bangladesh.

### 3.3 COVID-19 outbreak in the crop land, hilly areas and low land of Bangladesh

Bangladesh is known as a predominant agricultural country where more than 50% of the population rely directly or indirectly on employment in the agricultural and fisheries sectors, as indicated by the Food and Agricultural Organization of United Nation, 2020. These sectors are playing significant role in national economy by contributing 18% to country’s GDP as mentioned in the Bangladesh Economic Review. Thus, next we estimate the COVID-19 cases in several districts of Bangladesh that are highly associated with agricultural and fisheries sectors. The UKF-SIRD model estimates *∼* 1, 500 confirmed cases at the end of October for Noakhali district compared to ∼ 1, 000 cases for Patuakhali and Dinajpur region. In addition, haors (large shallow water body or backswamp) and wetlands area, such as Sunamganj has approximately ∼ 1, 200 cases, whereas Satkhira has ∼ 700 Corona virus positive cases in October, 2020. Notably, the model predicts lower number of confirmed cases (*<* 1, 000) and a smaller growth rate for high lands areas namely, Rangamati and Khagrachhari districts. Since these districts also have low *R*_*t*_ values (*<* 0.9) at the moment, there is a high chance that Rangamati and Khagrachhari will wipe out the COVID-19 outbreak if the current transmission trend continues for next 3-4 months. Taken together, this study represents that the COVID-19 pandemic is less serious in agricultural and high land areas compared to industrial areas in Bangladesh.

## Discussion

Epidemiological models are the powerful tools that are designed for the mathematical representations of the disease transmission and their associated dynamics [28, 29]. In addition, models can be used to estimate the trend of an epidemic outbreak under certain conditions so that necessary actions can be taken at various scales to restrain the disease diffusion in a community [30]. However, to make a series of policy decisions, it is a critical prerequisite to have a robust epidemiological model that is suitable for the intended purpose and shows accurate and precise prediction of a pandemic [31]. In this study, we integrated UKF with classic SIRD model and estimate COVID-19 diffusion in different parts of Bangladesh.

Dhaka is the 6^*t*^*h* largest densely populated city in the World, and the key financial hub which holds the major portions of the Bangladesh’s economy [32, 33]. Unfortunately, we observe that Dhaka is the epicenter of the COVID-19 pandemic with the highest number of infected cases due to its high population density in Bangladesh. Although the government has implemented zone-coded lockdown in small areas within the Dhaka city [34], we observe that the mobility between zones could not be maintained strictly due to economic activities. If the government had the capacity to test and isolate each positive cases, only then the zone-coded lockdown system in Bangladesh could have been successful. Several reports show the association of population density with higher rates of transmission, infection, and mortality from COVID-19 [35, 36]. Strikingly, our nationwide analysis of COVID-19 dynamics also demonstrate the relationship between population density and the COVID-19 diffusion rates for Bangladesh.

Industrial zones are at greater risk of transmitting the Corona virus as large groups of workers enter and leave the industrial area without maintaining social distance and proper hygiene in most of the cases. As a result, Narayanganj, Dhaka and Chattogram have been braving with COVID-19 for last couple of months. Notably, our model speculates the dire consequences for Chattogram if the COVID-19 diffusion continues with the current trend. Therefore, industrialists from Chattogram must ensure proper guideline for social distancing and the hygiene rules at the industrial areas to save their workers. Moreover, to reduce the public mobility, we suggest that public transportation between high risk areas should also be suspended for a time until the situation gets under control. To support this claim we would like to mention that the high rate of COVID-19 positive cases in the New York city of USA could have been controlled by implementing the proper lockdown and public mobility suspension [37].

According to our analysis, districts with the hilly areas and the wet land areas in Bangladesh show lower rate of infection than elsewhere. Besides the low population density in areas of Bandarban, Khagracchari and Rangamati, the government also control the transportation facilities in the area with some restrictions. On the other hand, the Cox’s Bazar, a popular tourist destination in Bangladesh has seen rise in COVID-19 positive cases despite having less population density. So it is evident that lower population density and restricted movements are the keys to slow down the diffusion rate in any areas. Therefore, we suggest to suspend or restrict the mass transportation between densely populated area, and industrial zone in a bit to contain the pandemic in Bangladesh.

Since Bangladesh is in emergency situation, we deal with incomplete and raw data of the current outbreak. For instance, if a person from a family becomes COVID-19 positive, the whole family might turn out to be positive. But, in many families only one of the members were tested for COVID-19 and if positive then counted as one despite the necessity to test for the entire family. Moreover, there were many cases who died with COVID-19 symptoms, but could not be tested, and conversely all the death cases counted in the analysis due to COVID-19 might have died by other diseases. In addition, the recovered cases count only considered those who went through two negative COVID-19 test results. Therefore, our UKF-SIRD model might not exemplify the pandemic situation accurately.

## Conclusion

Taken together, this study proposes a robust epidemiological method to assess the dynamics of COVID-19 pandemic in Bangladesh, as well as in any other countries which are still in the middle/late phase of the outbreak. Moreover, our estimation provides policymakers with a tool to evaluate the consequences of possible arrangements, for instance district wise lockdown, COVID-19 testing, social distancing and contact tracing. This data-driven mathematical modeling suggests that enforcing restricted social-distancing measures are obligatory and it needs to be maintained to contain the diffusion until the situation gets back to normal. We are optimistic that our UKF-SIRD model can serve as a base model for the policy suggestions and controlling the disease transmission in a heterogeneous region by adding more sophisticated factors under different environmental and political conditions in Bangladesh.

## Data Availability

All the data and relevant codebase is available at https://mjonyh.github.io.

## Supporting information

## Abbreviations

CFR: Case Fatality Rate
COVID-19: Coronavirus Disease 2019
IEDCR: Institute of Epidemiology, Disease Control and Research in Bangladesh
SIRD: Susceptible, Infected, Recovered and Death
UKF: Unscented Kalman Filter
WHO: World Heath Organization

## Declarations

### Ethics Approval and Consent to Participate

Not applicable.

### Consent for Publication

Not applicable.

### Author’s contributions

MH, MI, MA made substantial contributions to the conception or design of the work. MH and MI completed the relevant studies and created plots related to the SIRD estimation using Kalman filter, while MA critically revised the manuscript. All the authors met regularly to discuss the outcome of each experiment. The data were acquired from the IEDCR website and each of the results were cross validated by at least two of the three authors. All authors are responsible for acquiring, analysing, and interpreting the data for this article. All the authors approved the version to be published.

## Acknowledgements

We acknowledge the contribution of www.Pipilika.com software development team for collecting the daily data of total COVID-19 tests, total positive cases, and total deaths in Bangladesh.

